# Acute phase clinical manifestation of COVID-19 is linked to long-COVID symptoms; A 9-month follow-up study

**DOI:** 10.1101/2021.07.13.21260482

**Authors:** Fatemeh Sadat Mirfazeli, Atiye Sarabi-Jamab, Alireza kordi, Behnam Shariati, Seyed Vahid Shariat, Salar Bahrami, Shabnam Nohesara, Mostafa Almasi-Dooghaee, Seyed Hamid Reza Faiz

## Abstract

**Background:** The number of long-COVID is rising but it is not still clear which patients will develop long-covid and what will be the symptoms if they do.We followed up 95 patientswith confirmed COVID-19 after 9 months of the original study to delineate possible long COVID symptoms.

**Methods:** The original study included 201 patients who were treated in a large referral center from March to May 2020. Ninty percent of the patients reported physical or psychological symptoms within 9 months post-COVID.

**Findings:** Easy fatigability was the most common 51.04 % long-COVID symptoms followed by anxiety 38.54 %, dyspnea 38.54 %, and new headache 38.54%. There was no association between COVID-19 severity in the acute phase (admission status) and the number of long-COVID symptoms (F(1, 93) = 0.75, p = 0.45 (n.s.)), chronic fatigue syndrome (CFS) (F(1,93) = -0.49, p = 0.62 (n.s.), MOCA scores (F(1, 90) = 0.073, p = 0.787 (n.s.)) in the future. Being female (F(1, 92) = -2.27, p = 0.02), having a higher number of symptoms in the acute phase(F(1,93) = 2.76, p = 0.0068),and experiencing constitutional neuropsychiatric symptoms(F(1, 93)= 2.529, p = 0.01) in the acute phase were associated with higher occurance of CFS in follow up. Moreover, constitutional neuropsychiatric symptoms in acute phase were associated with cognitive dificits (lower MOCA score) (F(1, 93) = 10.84, p= 0.001) in the follow up.

**Conclusions:** Severity of the acute disease does not seem to be related to long-COVID symptoms. However, specific clinical presentations might be predictors of distinct long-COVID symptoms. Constitutional neuropsychiatric symptoms in the acute phase are associated with important and debilitating chronic symptoms including chronic fatigue syndrome, and cognitive deficits. These results might pave the way for findingthe underlying mechanisms of long-COVID and provide additional insight into possible candidate treatments for COVID-19.

## Introduction

As of this writing, more than 178 million cases of COVID-19 have been reported worldwide and more than 163 million have been recovered [1]. COVID-19 is recognized as a multi-organ disease with a broad spectrum of manifestations and there are increasing reports of prolonged effects after acute COVID-19 [2]. Some researchers define the term Post-Acute Sequelae of SARS-CoV-2 infection (PASC) for symptoms lasting more than 3 weeks after onset of the disease,and chronic symptoms of SARS-CoV-2 (CSSC) or long-COVID for those extending beyond 12 weeks [3, 4]. The prevalence of PASC and CSSCvary in different studies probably based on the severity of the acute symptoms, the duration of the follow-up, and other factors. In the study of Greenhalgh, T et al, it has beenreported 10% after 3 weeks [3]. In the post-acute COVID-19 US study performed 60 days after discharge, 6.7% of patients died,15.1% required readmission, and 32.6% of patients reported persistent or even new symptoms [5]. In another 6 months cohort study after discharge, excluding patients who were dead, readmitted,psychotic,demented, and immobile,76% of the patients had at least one residual symptom. In this study fatigue(63%), sleep difficulties(26%), hair loss (22%), and smell disorder(11%) were the most common residual symptoms [6]. Like several other respiratory viruses,COVID-19has neuroinvasive capacities. It may damage the CNS as a result of virus-induced neuro-immunopathology and/or virus-induced neuropathology. Other mechanisms of brain involvement may be due to the effect of the virus on other organs (such as the liver, kidneys, and hypoxia) and virus-induced coagulopathy. The psychosocial effects of the disease, such asisolation and distress about the disease and its consequences are other important issues after discharge[7]. Therefore, it is not surprising that neuropsychiatric symptoms are among the most common Post-Acutesymptoms. In one studyover 30% of hospitalized COVID-19patients exhibited cognitive problems, depression, and anxiety which persisted for months after discharge [7]. One month followinginfection there was a considerably high prevalence of insomnia (34-40%), depressivesymptoms (31-45%), anxiety symptoms (22-47%), Post Traumatic Stress Disorder (28%), and obsessive-compulsive symptoms (20%)[8, 9]. In a recent survey, Perils et al. found that 52.4% of COVID-19 patients had depressive symptoms according to Patient Health Questionnaire–9 (PHQ-9). In their study presence of headache was associated with greater probability of moderate or more severe depressive symptoms[10].

Neurocognitive symptoms are another category of post-COVID symptoms. In one study,34% of 279 hospitalized COVID-19 patients, reported memory loss and 28% described impaired concentration three months after discharge[11]. Other cognitive symptoms including impaired memory and receptive language and/or executive dysfunction have been reported[12]. New-onset dementia after hospitalization for COVID-19 was reported 2–3 times more common than what was observed after hospitalization for other medical conditions[13]. However, it is not yet clear which patients will develop PACS and CSSC symptoms, specifically chronic neuropsychiatric symptoms, after infection with COVID-19. A few efforts have been performed to find the association between clinical manifestations and sociodemographic factors of patients and neurocognitive sequelae of the COVID-19 disease in the future.For example, Woo,MS et al. studied 18 patients with mild to moderate COVID□19 disease with a mean age of 42 years and found that after around 85 days, over 75% of patients had difficulties in episodic memory, attention, and concentration,but these cognitive deficits were not associated with a history of fatigue, depression, hospitalization, treatment, viremia or acute inflammation[14]. However,in other studies, delirium as one of the most important neurocognitive symptoms during the acute phase of illness was suggested to contribute to long-term cognitive decline[15, 16]. Moreover, in other studies, the post-COVID manifestations were reported to be related to comorbidities and disease severity[17]. Most of the literature on predictors of long-COVID symptoms is whether subject to small sample size[14] or controversial results[15-17]. Therefore in this study, we tried to investigate the association of acute clinical manifestation of the COVID-19 and sociodemographic features of the patients, and the experience of long-COVID symptoms.We aimed to follow the patients we studied 9 months ago[18]. Neuropsychiatric symptoms were grouped into 3 distinct categories:anosmia and hypogeusia (olfactory symptoms); dizziness, headache, and limb force reduction (general constitutional-like neuropsychiatric symptoms); photophobia, mental state change, hallucination, vision and speech problem, seizure, stroke, and balance disturbance (specific neuropsychiatric symptoms or specific CNS type). There were also three non-neuropsychiatric clusters of symptoms: diarrhea and nausea (gastrointestinal symptoms); cough and dyspnea (respiratory symptoms); and fever and weakness (constitutional symptoms).The cluster of GI symptoms was closer to the neurological symptoms (specifically the ones with olfactory and general constitutional-like symptoms) [18].

## Methods

### Participants

The present study was a 9-month follow-up of 201 participants with a diagnosis of COVID-19 (52 outpatients and 149 inpatients)from March 2019 to April 2020 in a large referral Hospital in Tehran (Rasool Akram Medical Complex). The diagnosis was based on a positive real-time reversetranscription-polymerase chain reaction assay using a COVID-19nucleic acid detection kit or atypical chest CT scan for COVID-19 reported by three specialists (an emergency medicine specialist, an infectious disease specialist, and a pulmonologist). The diagnosis was supported by related laboratory tests (biochemistry andinflammatory factors). Patients were diagnosed with COVID-19 according to WHO interim guidelines[19].

A total of 94 from 201 patients (median age = 50 [range 28 -86], 58% male [n = 55]) were responsive. We follow Patients at a median of 270 days (range=258-300). Eight patients passed away in the hospital or a couple of months following their COVID-19 diagnosis.

All of the patients gave informed consent for the study. This research hasbeen approved by the ethics committee of the Iran University of Medical Science (code number: IR.IUMS.REC.1399.1040).

### Data collection

In the originalstudy [18], patients with mild to moderate COVID-19 filled out an online comprehensive symptom checklist and a trained neurologic resident examined patients with moderate to severe who were hospitalized. Patients’ electronicparaclinical data such as imaging and laboratory tests were also recruited.

In the current study, two trained medical studentsfollowed up with the patients via phone interview. The data about the clinical and paraclinical manifestation of the patients, drug history, comorbidity,sociodemographic profile that we collected 9 months before in the first study was also collected.

### Symptom report

We documented symptoms reported after recovery of COVID-19 using a comprehensive checklist of specific symptoms of different organs potentially correlated with COVID-19, drug history, demographic information, comorbidity. We also registered chronic fatigue syndrome based on CDC criteria[20],which is a neurological disease characterized by severe mental and physical fatigue, chronic pain, and sleep disorder. Patients were asked to specifically remember symptoms that started after their diagnosis of COVID-19, those that started after recovery from COVID-19, persisted for a long time.

### Cognitive assessment

We used a Persian translated version of Montreal Cognitive Assessment (MoCA)-BLIND 7.1 for telephone cognitive assessment. The MOCA is extensively used for mildly neurocognitive impairment screening [21]. After exclusion of its visual component, it has been validated for cognitive assessment of the patients via telephone follow-up [22-24]

### Analysis

All statistical analyses were performed using R (version 3.6.1; R Core Team, 2019) in RStudio (RStudio-1.2.5001). For numerical variables, we used mean and standard deviation (SD) when they were normally distributed and we used range if they were not. Continuous variables were compared using the Wilcoxon rank-sum test. To find the predictor factor in our laboratory measures, we performed a (generalized) linear model and in the case of more than one predictor factor, we performed the stepwise regression by iteratively adding and removing variables in each step. To do this in R, we used the “MASS” package, which chooses the best model based on the AIC criterion, and we used the “stepAIC” function for stepwise regression. The significance threshold at a 2-sided P value was set at less than 0.05.

## Results

In the descriptive analysis easy fatigability 51.04%, was found to be the most common long COVID symptom followed by anxiety 38.54%, dyspnea 38.54%, and new headache 38.54%.Eighty-six (90%) patients showed at least onelong-COVID symptom in their follow-up. figure 1.

**Figure 1:**
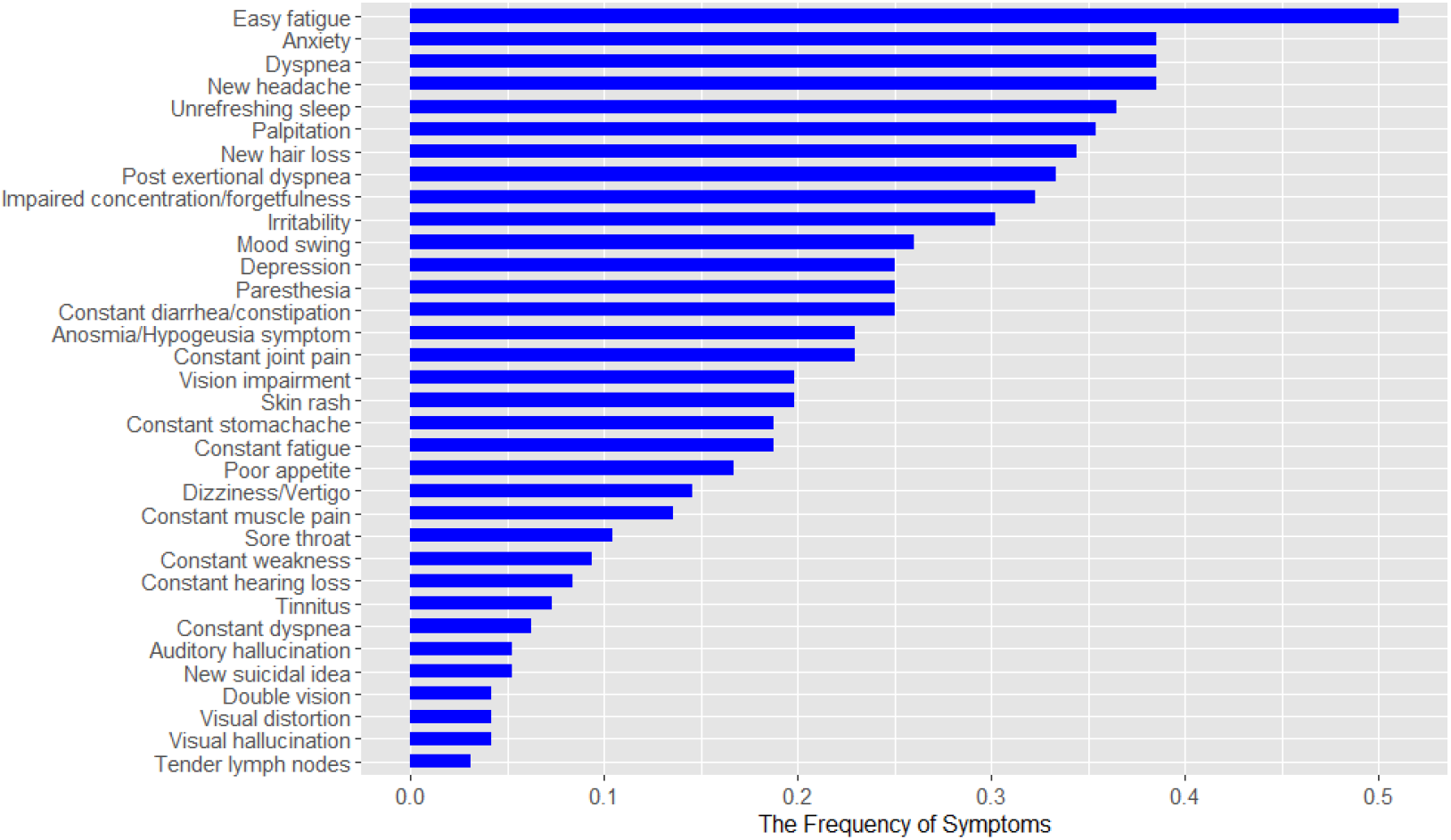
The frequency of long-COVID symptoms in the follow-up of 95 patients with history of COVID-19.

The persistent symptoms were then clustered based on similarity of symptoms and the symptoms clustered in the acute phase in the previous study. As you can see there were 8 main clusters: constitutional neuropsychiatric symptoms (including: Palpitation, Dizziness/Vertigo, Anxiety, Moodswing, Irritability, Newsuicidalidea, Paresthesia, Depression), dermatologic symptoms(including: Skinrash, Newhairloss), respiratory symptoms (including: Postexertionaldyspnea, Constantdyspnea, Dyspnea), GI symptoms (including: Constant.diarrhea/constipation, Constant.stomach.ache, Poor.appetite), Chronic fatigue syndrome (including: Sore throat, New headache, Constant musclepain, Constant joint pain, Easy.fatigue, Constant.fatigue, Constantweakness, Impairedconcentration/forgetfulness, Unrefreshing sleep, Tenderlymphnodes), specific neuropsychiatric symptoms (including: Auditoryhallucination, Visualhallucination, Visionimpairment, Visualdistortion, Doublevision), ENT symptoms (including: Constanthearingloss, Tinnitus). Constitutional neuropsychiatric symptoms are the most common reported cluster of symptoms 58.33% .figure 2.

**Figure 2:**
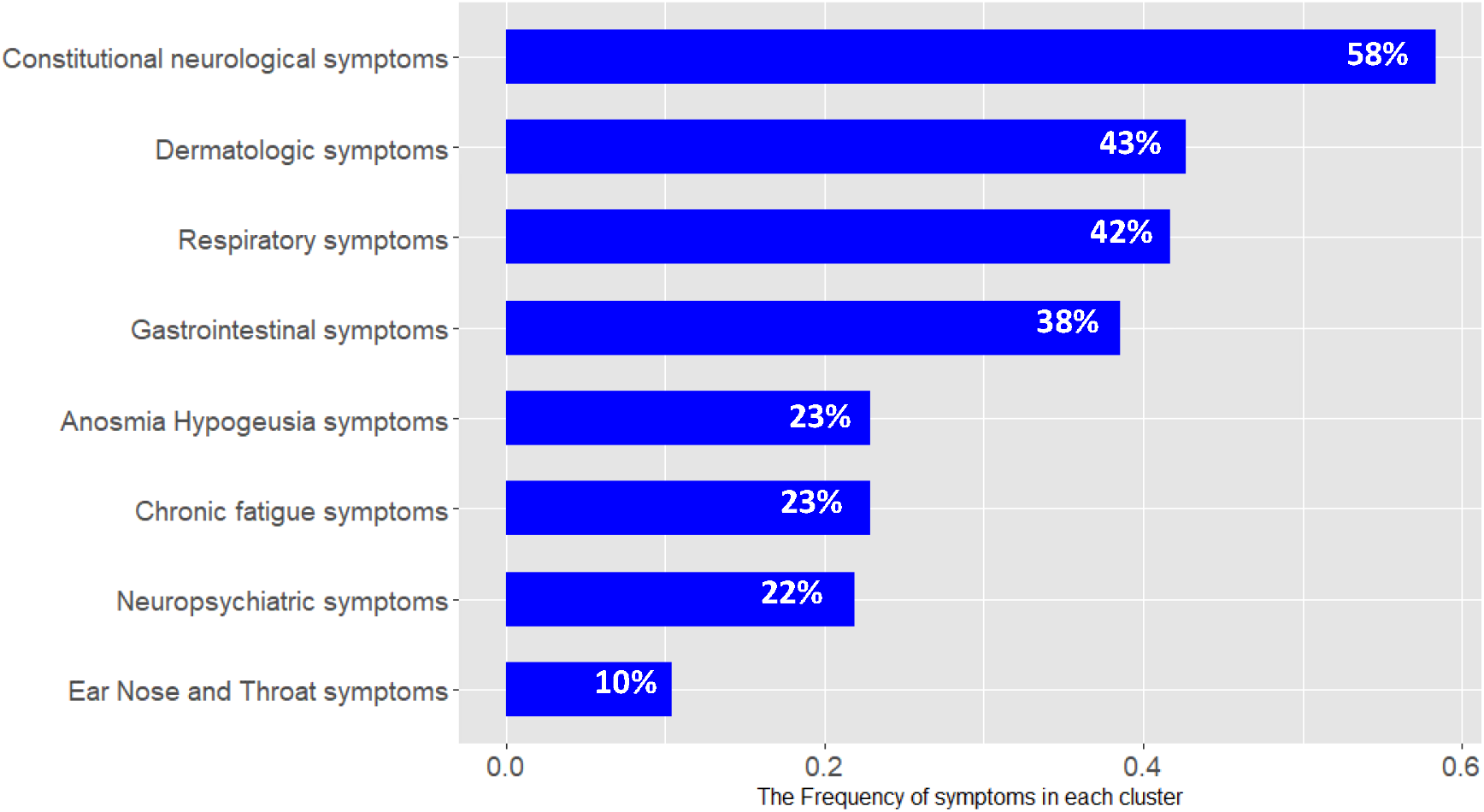
The frequency of clusters of long-COVID symptoms in the follow-up of 95 patients with history ofCOVID-19.

Linear regression analysisin exploring sociodemographic predictors of chronic fatigue syndrome indicated that gender was a predictor of chronic fatigue syndrome, as this syndrome was more common in the female patients: 64% Female (median age = 49 [range 36 -86]) 36% Male (median age = 48.5 [range 39 -77];(F(1, 92) = -2.27, p = 0.02), while age was not(F(1, 92) = 1.27, p = 0.2 (n. s.)). Figure 3.

**Figure 3.**
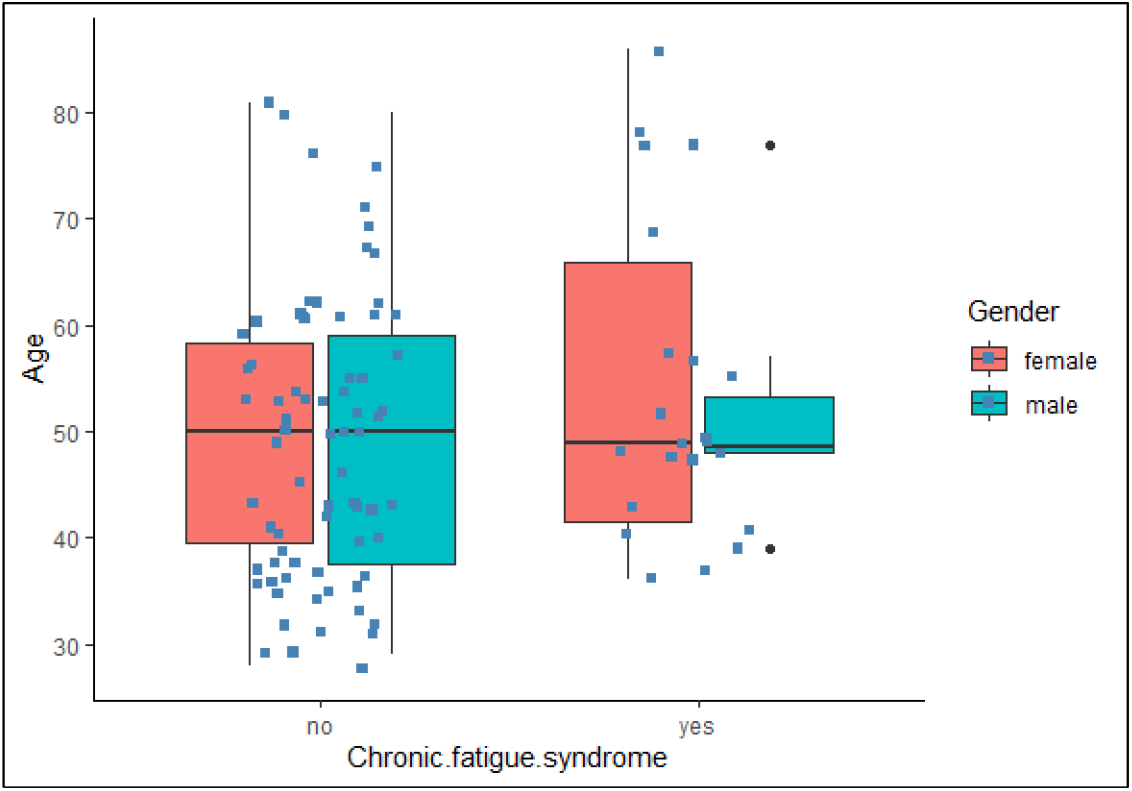
The relation of chronic fatigue syndrome (CFS) and age/ gender.

### The relation between clinical manifestations in the acute phase of COVID-19 and long COVID symptoms

In our previous study[18], we labeled the patients’ symptoms in six clusters: Constitutional-like Neuropsychiatric cluster (including dizziness, headache, and limb force reduction symptoms), Olfactory cluster(anosmia and hypogeusia symptoms), Constitutional cluster (including fever and weakness symptoms), Neuropsychiatric cluster (including photophobia, mental state change, hallucination, vision and speech problem, seizure, stroke, and balance disturbance symptoms), GI cluster(including diarrhea and nausea symptoms), and Respiratory cluster (including cough and dyspnea symptoms). Then, it was explored how the new persistent cluster symptoms reported by patients (long-COVID), which were classified in eight different classes in the previous section, are associated with early symptoms categories.To this end, multiple regression analysis was used to test if the early COVID symptoms could predict each of the new clusters or symptoms preserving within 9 months. The result of the general linear regression model indicated that the constitutional neuropsychiatric symptoms in the acute phase predictedthe chronic fatigue syndromein the future as long-COVID symptoms (R^2^ =6.4%F(1, 93)= 2.529, p= 0.01).Further regression analysis showed that specific neuropsychiatric symptoms in the acute phase are associated with GI symptoms (R^2^ = 6%, F(1,93)= 2.43, p = 0.01)in the future. Having specific neuropsychiatric symptoms in the acute phase is also a predictor of dermatologic disturbances in the future (R^2^ = 8.5%, F(1,93)= 2.96, p = 0.003). Moreover, specific neuropsychiatric symptomspredicted the respiratory symptoms as long-COVID (R^2^= 9.8%, F(1,93)=3.19, p = 0.001). Respiratory symptoms in the acute phase predicted anosmia and dysgeusia symptoms in the future. They also marginally predictedolfactory symptoms in the acute phase of COVID-19, and finally those with constitutional and olfactory symptoms, in the acute phase of COVID-19,were predictors of ENT symptoms as long-COVID (R^2^=16%, F(2,93)=-1.913, p = 0.5, F(2,93)= 3.62, p = 0.0004 respectively).

There was no association between COVID-19 severity in the acute phase (admission status) and the number of long-COVID symptoms(F(1, 93) = 0.75, p = 0.45 (n.s.)). It was also no association between COVID-19 severity in the acute phase (admission status) and having CFS in the future as a long-COVID symptom (F(1,93) = -0.49, p = 0.62 (n.s.). However, the number of symptoms in the acute phase could predict having CFS in the future as long-COVID(F(1,93) = 2.76, p = 0.0068), Figure 4.

**Figure 4.**
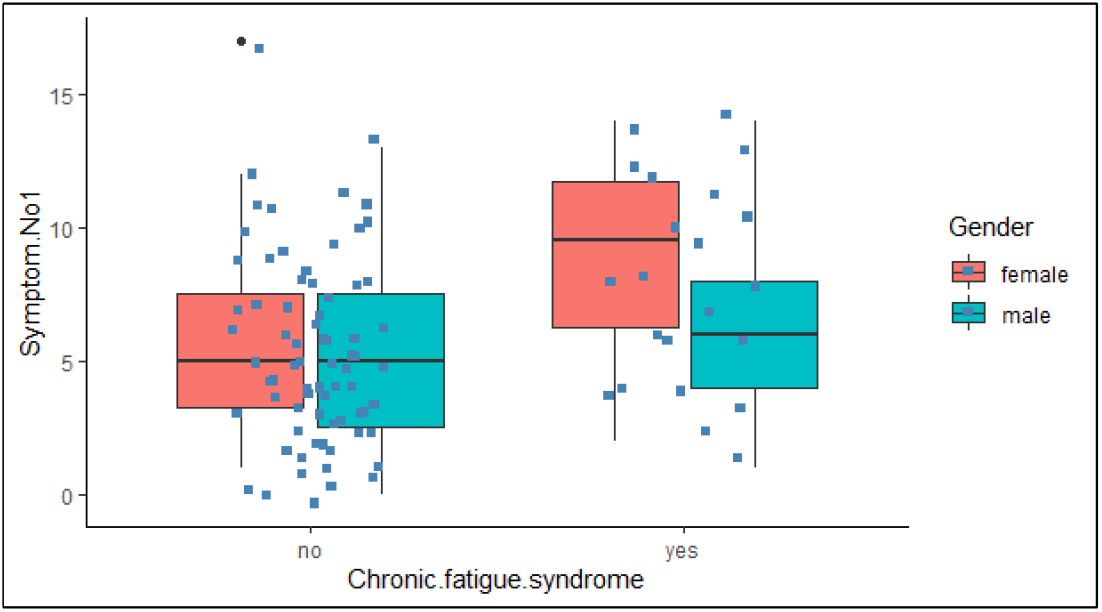
The relation of chronic fatigue syndrome (CFS) and the number of symptoms in the acute phase.

**Figure 5.**
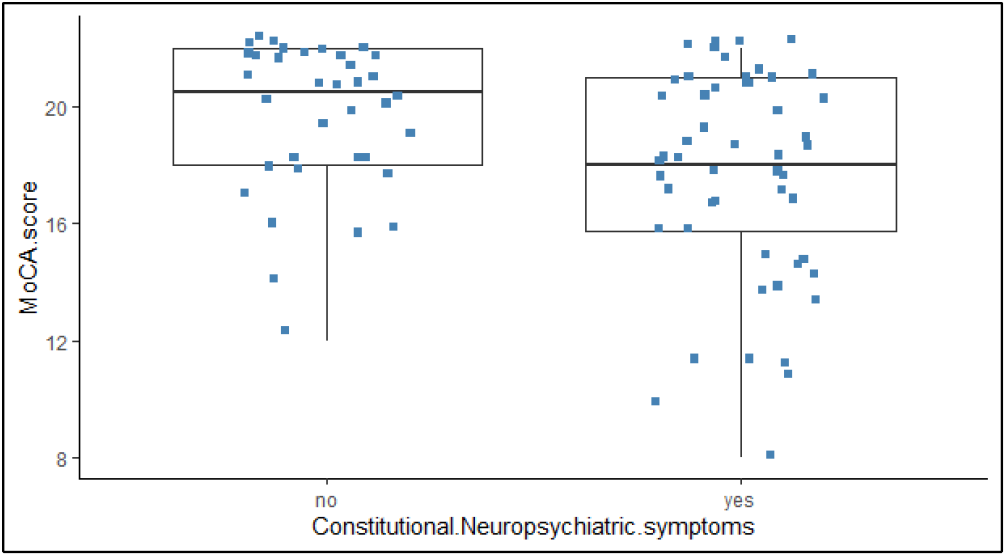
The relation between the constitutional neuropsychiatric symptoms and follow-up MOCA score.

### Cognitive status in the follow-up of patients with COVID-19

To know about the relation between MOCA score and severity of COVID-19, Regression analysis did not show any correlation between MOCA score in the follow-up (cognitive status) and disease severity (admission history) of patients with COVID-19 (F(1, 90) = 0.073, p = 0.787 (n.s.)). However, it was shown that the duration of COVID-19is associated with a lower MOCA score but it was confounded by the effect of age. (F(1,86)= 2.54, p = 0.11, F(2,85) = 26.04, p = 1.5e-9).Furthermore, the number of symptoms in the acute phase also did not predict the MOCA score in the future (R^2^ = 6.8e-5 %, F(1,92)= 0.079, p = 0.937 (n.s.)).

Moreover, ANOVA analysis showed that among six symptoms categories at the acute phase of COVID-19,those with constitutional neuropsychiatric symptoms reported lower MOCA scores (F(1, 93) = 7.17, p= 0.008) and had poor performance on delayed recall (F(1, 93) = 8.91, p= 0.003).

## Discussion

Our study was one of the few attempts to follow-up the patients within 9 months course after registering and clustering clinical manifestation of COVID-19 in the acute phase[18]. We found that there was an association between acute phase clinical manifestations of COVID-19 and long-COVID symptoms and cognitive sequelae in the future. However, we did not find any association between the severity of symptoms in the acute phase and long-COVID symptoms and cognitive sequelae in the future.

Long-COVID symptoms were very common among our patients. 90% of our patients reported physical or psychological symptoms within 9 months post-COVID. Easy fatigability 51.04 %, was found to be the most common long-COVID symptoms followed by anxiety 38.54 %, dyspnea 38.54 %, and new headache 38.54%. Constitutional neuropsychiatric symptoms were the most common cluster of symptoms 58.33% reported within the 9-month follow-up. Chronic fatigue syndrome was reported in 22.92% of patients. Our study is consistent with previous studies that reported a high prevalence of long-COVID symptoms in follow-ups [25]. For example, Sykes et al reported long-COVID symptoms in eighty-six percent of their follow-up patients[25]. Consistent with other studies[26], there was no association between the disease severity in the acute phase of COVID-19 and the risk of developing long-COVID in the future. However, we showed that these post-acute symptoms could last quite for a long time, up to 9 months while previous literature mainly followed COVID-19 patients for a maximumof 6 months[2, 25, 27]. Experiencing physical symptoms along with psychological and cognitive symptomsfor several months couldput a burden on people with a history of COVID-19. Moreover, having psychological, cognitive, and physical symptoms would also suggest a biopsychosocial underlying mechanism for long-COVID, not merely apsychological underlying mechanism. Some studies implied that long-COVID represents a miscellany of physical and psychological symptoms, like Persian gulf war illnessand it could be a post-traumatic syndrome[25]. However, we found that there wasan association between the symptom categories of acute clinical manifestation of COVID-19 and long-COVID symptoms and they were not just random miscellany of symptoms coming together which makes long-COVID as just a post-traumatic stress syndromeless probable[28]. We found that constitutional neuropsychiatric symptom categories were associated with having a lower MOCA score in the future; while the number of COVID-19 symptoms in the acute phase, the severity of COVID-19 presented by the status of admission, and the duration of COVID-19 symptoms in the acute phase were not. These findings show that the pattern of organ involvement in COVID-19 specifically in the acute phase would show how you would experience your long-COVID in the future. It seems that systemic inflammation, even when it is not severe could lead to cognitive impairment in the future if it involves neuroinvasion in the acute phase [29, 30]. Our findings were consistent with previous studies which reported an age-dependent cognitive decline in the follow-up of COVID-19 patients[31]. Among MOCA scores, we found that impaired delayed recall was mostly associated with having constitutional neuropsychiatric symptoms in the acute phase. In other studies, different cognitive sequelae such as apathy, executive dysfunction, reduction in global cognition were reported and GABAergic dysbalance have been proposed as the underlying mechanism for executive dysfunction [32].

We also found that neuropsychiatric involvement other than a cognitive decline in the acute phase was a good predictor of long-COVID in different organs in the future. Experiencing specific neuropsychiatric symptom categories in the acute phase was associated with specific GI, dermatologic, and respiratory symptoms in the future. Martina Sollina et al, in their study, showed that vascular inflammation could be the leading cause of long-lasting COVID symptoms [28]. It is not yet clear whether these organs share a similar receptor in their vasculature or other cellular structure.

We also found that those with respiratory symptomsin the acute phase were associated with longer anosmia and dysgeusia problems. So it seems that not all of the infected individuals would suffer from long olfactory problems, but it is more probable in those who experience COVID-19 with lung involvement.

Those with constitutional and olfactory symptoms, in the acute phase of COVID-19, were associated with ENT (tinnitus and hearing loss) symptoms in the future.

All of these could be explained by similar receptors in the vascular structure of different organs as a result of systemic inflammatory responses [28]. It could be a matter of future debate and exploration.

Being female, experiencingthe constitutional neuropsychiatric symptom categories in the acute phase, and having a higher number of symptoms in the acute phase could predict CFS in the future. This was consistent withother results in our study that the severity of the COVID -19 presented by admission status in the acute phase could not predict developing long-COVID in the future. It seems that it is also true for chronic fatigue syndrome and what is more important to predict the risk of CFS in the future is the pattern of organ involvement in the acute phase rather than the severity of the disease (low saturation, loss of consciousness, shock, admission status). It would suggest the mechanism of CFS is similar to other long-COVID symptoms. These findings are in line with previous studies [33]. Liam Townsend et al., also found that COVID persistent fatigue to be more common in females and is not associated with acute illness severity (admission status and laboratory inflammatory markers) [33]. One possible mechanism would be a state of chronic low-grade neuroinflammation followed by the disease [29]. COVID-19 can infect the brain leading to neuroinflammation [34]. Alternatively, it can cause inflammation in another part of the body, activating an innate immune response in the brain via humoral response and retrograde vagus nerve travel [30, 35]. Neuroinflammation can cause severe fatigue [36]. This will explain the association of constitutional neuropsychiatric symptoms in the acute phase and CFS in the future.

All in all, finding both physical and psychological long-COVID symptoms emphasizes the need for a multidisciplinary rehabilitation team in COVID-19 survivors.

However, our study was not without limitations. A number of patients from the original study who did not cooperate in the second study could change the current result. A small number of cases had full workup of paraclinical data and chest imaging in the electronic system of the hospital from the original study; therefore, finding an association between blood marker and long-COVID symptoms was not possible. Future studies to explore the association between symptom categories and inflammatory markers in the acute phase and long-COVID symptoms could shed new light on the underlying mechanism of the post-acute phase in people suffering from COVID-19.

## Conclusion

Most of our patients experienced at least one physical, psychological or cognitive symptoms within 9 months following COVID-19. Easy fatigability, followed by headache, anxiety, and dyspnea were the most common long-COVID symptoms. There was an association between the acute presentation of COVID-19 and long-COVID symptoms in the follow-up, which was not related to disease severity. Having constitutional neuropsychiatric symptoms in the acute phase was associated with chronic fatigue syndrome, lower MOCA score, and poorer performance on delayed recall in future. Having specific neuropsychiatric symptoms in the acute phase was associated with dermatologic, GI, and respiratory symptoms in the future as long-COVID. These results might pave the way for finding the underlying mechanisms oflong-COVID and provide additional insight into possible candidate treatmentsfor COVID-19.

## Data Availability

Data are available upon reasonable request from corresponding author.

## Declarations

### Ethics approval and consent to participate

This research hasbeen approved by the ethics committee of the Iran University of Medical Science (code number: IR.IUMS.REC.1399.1040).

### Informed consent

All patients included in the study signed informed consent for their participation in the study. All patients’ information remained confidential.

## Acknowledgment

The authors would like to thank the Rasool Akram Medical Complex Clinical Research Development Center (RCRDC) for its technical and editorial assists.

## Competing interests

All authors declare that they have no conflict of interest to disclose.

## Funding

The authors received no financial support for the research, authorship, and/or publication of this article.

## Availability of data and materials

The datasets used and analysed during the current study are available from the corresponding author on reasonable request.

## Authors’ contributions

FSM, and ASJ contributed as first authors.

FSM, and ASJ: conceptualization and design of the manuscript; Ak, BS, SVS and SHRF: drafting of the manuscript; Ak, BS, SVS, SB, SN, MAD: reviewing the literature and writing the manuscript. Ak, BS, and SVS: the statistical analysis and methodology. SB, SN, MAD acquisition of the data; FSM, ASJ and SHRF were supervisions. All the authors: revising the paper for critically important intellectual content, and final approval of the version to be published; All authors contributed to editing the manuscript. The corresponding author attests that all listed authors meet authorship criteria and that no others meeting the criteria have been omitted. SHRF is guarantor.

## Notes

### Competing Interest Statement

The authors have declared no competing interest.

### Funding Statement

No funding

### Author Declarations

Ethics approval and consent to participate This research has been approved by the ethics committee of the Iran University of Medical Science (code number: IR.IUMS.REC.1399.1040).

